# Performance of Abbott Architect, Ortho Vitros, and Euroimmun Assays in Detecting Prior SARS-CoV-2 Infection

**DOI:** 10.1101/2020.07.29.20164343

**Authors:** Shiwani Mahajan, Carrie A. Redlich, Adam V. Wisnewski, Louis E. Fazen, Lokinendi V. Rao, Karthik Kuppusamy, Albert I. Ko, Harlan M. Krumholz

**Affiliations:** Center for Outcomes Research and Evaluation, Yale-New Haven Hospital, New Haven, Connecticut; Section of Cardiovascular Medicine, Department of Internal Medicine, Yale School of Medicine, New Haven, Connecticut; Yale Occupational and Environmental Medicine Program, Department of Internal Medicine, Yale School of Medicine, New Haven, Connecticut; Department of Pathology, University of Massachusetts Medical School, Worcester, Massachusetts; Quest Diagnostics, Marlborough, Massachusetts; Department of Epidemiology of Microbial Diseases, Yale School of Public Health, New Haven, Connecticut; Department of Health Policy and Management, Yale School of Public Health, New Haven, Connecticut

## Abstract

**Background:** Several serological assays have been developed to detect anti-SARS-CoV-2 IgG antibodies, but evidence about their comparative performance is limited. We sought to assess the sensitivity of four anti-SARS-CoV-2 IgG enzyme-linked immunosorbent assays (ELISA) in individuals with evidence of prior SARS-CoV-2 infection.

**Methods:** We obtained sera from 36 individuals with PCR-confirmed SARS-CoV-2 infection between March and May 2020. We evaluated samples collected at around 21 days (±14 days) after their initial PCR test using 3 commercially available ELISA assays, two anti-spike (Ortho-Clinical Diagnostics Vitros, and Euroimmun) and one anti-nucleocapsid (Abbott Architect), and a Yale-developed anti-spike ELISA test. We determined the sensitivity of the tests and compared their results. The Euroimmun and Yale ELISA had an equivocal and indeterminate category, which were considered as both negative and positive.

**Results:** Among the 36 individuals with SARS-CoV-2 infection, mean age was 43 (±13) years and 19 (53%) were female. The sensitivities of the tests were not significantly different (Abbott Architect, Ortho Vitros, Euroimmmun, and Yale assays: 86% (95% confidence interval [CI], 71– 95), 94% (95% CI, 81–99), 86% (95% CI, 71–95), and 94% (95% CI, 81–99), respectively; p- value=0.464). The sensitivities of the Euroimmun and Yale ELISA tests increased when the equivocal/indeterminate results were considered positive (97% [95% CI, 85–100] and 100% [95% CI, 90–100], respectively), but were not significantly different from other tests (p=0.082). The cross-correlation coefficient ranged from 0.85–0.98 between three anti-spike protein assays (Ortho Vitros, Euroimmun, Yale) and was 0.58–0.71 between the three anti-spike protein assays and the anti-nucleocapsid assay (Abbott).

**Conclusion:** The sensitivities of four anti-SARS-CoV-2 protein assays did not significantly differ, although the sample size was small. Sensitivity also depended on the interpretation of equivocal and indeterminate results. The strongest correlations were present for the three anti-spike proteins assays. These findings suggest that individual test characteristics and the correlation between different tests should be considered when comparing or aggregating data across different populations studies for serologic surveillance of past SARS-CoV-2 infection.

## BACKGROUND

Serological testing is being increasingly used to evaluate people for antibodies to severe acute respiratory syndrome coronavirus 2 (SARS-CoV-2). Recently, a number of serology tests have been developed and have received Food and Drug Administration (FDA) Emergency Use Authorization (EUA) for the detection of these antibodies.^1^ However, there are relatively few evaluations of these tests from a cross-section of individuals with polymerase chain reaction test confirmed SARS-CoV-2 infection.

Accordingly, we sought to determine the sensitivity of three commercially available SARS-CoV-2 IgG assays and an anti-spike enzyme-linked immunosorbent assay (ELISA) test at Yale University, in preparation for conducting a large seroprevalence survey of a representative population sample for the state of Connecticut, USA. We compared the Abbott Architect, the Ortho Vitros and Euroimmun, all of which are approved by the US FDA. The Abbott Architect SARS-CoV-2 IgG assay detects antibodies against the nucleocapsid, whereas the Ortho Vitros anti-SARS-CoV-2 IgG test and Euroimmun SARS-CoV-2 ELISA detect antibodies against the spike protein of the virus.^2^ We also compared the results of these antibody tests with each other.

## MATERIAL AND METHODS

### Patient Sample

We took advantage of previously collected and de-identified serum specimens from 36 individuals with confirmed SARS-CoV-2 infection between March and May 2020 and the availability of resources to test them with the commercially available assays, which set the sample size. Patients were defined as SARS-CoV-2 positive based on reverse transcription polymerase chain reaction (RT-PCR) testing on nasopharyngeal swab specimens collected. The serology samples included individuals with variable severity and duration of symptoms, but all samples were collected when individuals were asymptomatic and in convalescence, approximately 21 days (±14 days) after their initial RT-PCR test. For non-hospitalized individuals, answers to questions from survey data and telephone interviews were used to identify individuals who self-reported symptoms that they attributed to COVID-19 infection. All individuals provided informed consent and samples were collected in accordance with protocols approved by the Institutional Review Board at Yale University.

### Sampling Methodology and IgG Testing

Blood was collected in vacutainer tubes, and serum was separated. Serum aliquots were stored at -80°C. Samples had previously been assayed on a laboratory developed test at Yale, an ELISA against the spike antigen, but samples were selected blinded from these results. A cutoff of 0.32 was used for the Yale anti-spike ELISA to identify positive samples, with 0.28-0.32 considered indeterminate. Approximately 450uL of serum from each of the 36 samples was sent from Yale to Quest Diagnostics for testing on their 3 commercially-available SARS-CoV-2 IgG assays, namely – Abbott Architect SARS-CoV-2 IgG test, Ortho-Clinical Diagnostics Vitros anti-SARS-CoV-2 IgG test, and Euroimmun SARS-CoV-2 ELISA (IgG).^2^ Testing was performed in a blinded manner.

Antibody levels were expressed as the ratio of chemiluminescence values over the cutoff (S/CO) values. The following cutoffs were used for reporting of results according to the manufacturer’s instructions: 1) Abbott Architect IgG assay^3^: <1.4 = negative and ≥1.4 = positive; 2) Ortho Vitros IgG assay^4^: <1.00 = negative and ≥1.00 = positive; and 3) Euroimmun IgG assay^5^: <0.8 = negative, ≥0.8 to <1.1 = equivocal, and ≥1.1 = positive.

### Data Analysis and Visualization

We collected data on the optical density of the Yale anti-spike ELISA and the index values for Abbott Architect, Ortho Vitros, and Euroimmun SARS-CoV-2 IgG assays, and compared the results reported by the four serology assays. The sensitivity of different assays was calculated, and Fishers exact test was used to detect for statistical difference. The correlation between the index values of different assays and optical density of Yale anti-spike ELISA was assessed using Spearman’s correlation coefficient, with confidence intervals (CI) calculated by z-transformation. Individual scatter plots were created for each two-way test comparison with non-parametric locally weighted regression (Loess) smoothing with 95% CI. SAS and R (version 3.6.1) was used for all statistical analyses and graphical and tabular preparations. A p-value of <0.05 was considered statistically significant.

## RESULTS

### Patient Characteristics

Of the 36 individuals with SARS-CoV-2 infection included in the study, mean age 43 (±13) years and 19 were female, 16 were male, and 1 did not have information available on gender.

### Sensitivity of the SARS-CoV-2 IgG Assays

Sensitivity of the Abbott Architect, Ortho Vitros, Euroimmun, and Yale ELISA SARS-CoV-2 IgG assays is shown in **Table 1**, based on the results in 36 individuals with RT-PCR confirmed SARS-CoV-2 infection.

**Table 1.** Sensitivity of different SARS-CoV-2 IgG Assays (n = 36 positive RT-PCR)

| **SARS-CoV-2 IgG Assay** | **True Positives, n** | **False Negatives, n** | **Sensitivity, % (95% CI)** |
| --- | --- | --- | --- |
| Abbott Architect | 31 | 5 | 86% (71% to 95%) |
| Vitros | 34 | 2 | 94% (81% to 99%) |
| Euroimmun |  |  |  |
| Equivocal results as positive | 35 | 1 | 97% (85% to 99%) |
| Equivocal results as negative | 31 | 5 | 86% (71% to 95%) |
| Yale ELISA |  |  |  |
| Indeterminate as positives | 36 | 0 | 100% (90% to 100%) |
| Indeterminate as negative | 34 | 2 | 94% (81% to 99%) |
| Abbreviations: CI, confidence interval; ELISA, enzyme-linked immunosorbent assay; SARS-CoV-2, severe acute respiratory syndrome coronavirus 2. |  |  |  |

The Abbott Architect IgG assay identified 31 samples as true positives and 5 samples as false negatives, and had a sensitivity of 86% (95% CI, 71% to 95%). The Othro Vitros IgG assay identified 34 samples as true positives and 2 samples as false negatives, yielding a sensitivity of 94% (95% CI, 81% to 99%). Euroimmun IgG assay identified 31 samples as true positives, 1 sample as false negative, and 4 as equivocal. The sensitivity of the assay considering equivocal samples as positives was 97% (95% CI, 85% to 100%), whereas the sensitivity was 86% (95% CI, 71% to 95%) when the equivocal samples were considered as negatives. The Yale anti-spike ELISA identified 34 samples as true positives and 2 as indeterminate. The sensitivity of the assay considering the indeterminate as positives was 100% (95% CI, 90% to 100%), whereas the sensitivity was 94% (95% CI, 91% to 99%) when the indeterminate samples were considered as negatives. Overall, sensitivities of the four anti-SARS-CoV-2 protein assays did not differ significantly whether indeterminate/equivocal samples were considered as positives (p=0.082) or as negatives (p=0.464).

### Correlation Between Different Assays

The 36 serum samples from individuals with RT-PCR-confirmed SARS-CoV-2 infection showed varied index values on Abbott (range 0.02–8.81), Ortho Vitros (range 0.01–20.70), and Euroimmun (range 0.13–8.60) IgG assays. The optical density of IgG on the Yale anti-spike ELISA ranged from 0.30 to 3.07.

The index values of Abbott Architect, Ortho Vitros, and Euroimmun SARS-CoV-2 IgG assays had a correlation of 0.58 (95% CI, 0.32 to 0.77), 0.85 (95% CI, 0.72 to 0.92), and 0.85 (95% CI, 0.72 to 0.92), respectively, compared with the optical density of Yale anti-spike ELISA (**Figure 1 A, B, C)**. The correlations between the index values of the 3 different commercial SARS-CoV-2 IgG assays are shown in **Figure 1 (D, E, F)**. The Ortho Vitros and Abbott Architect SARS-CoV-2 IgG assays had a correlation of 0.71 (95% CI, 0.50 to 0.84), Euroimmun and Abbott Architect IgG assays had a correlation of 0.66 (95% CI, 0.42 to 0.81), and Euroimmun and Ortho Vitros IgG assays had a correlation of 0.98 (95% CI, 0.96 to 0.99). Compared with each other and with the Yale anti-spike ELISA, the Euroimmun and Ortho Vitros IgG assays had the highest correlation, and Abbott Architect assay had the lowest correlations when compared with the other two assays, as well as to the Yale anti-spike ELISA.

**Figure 1.**
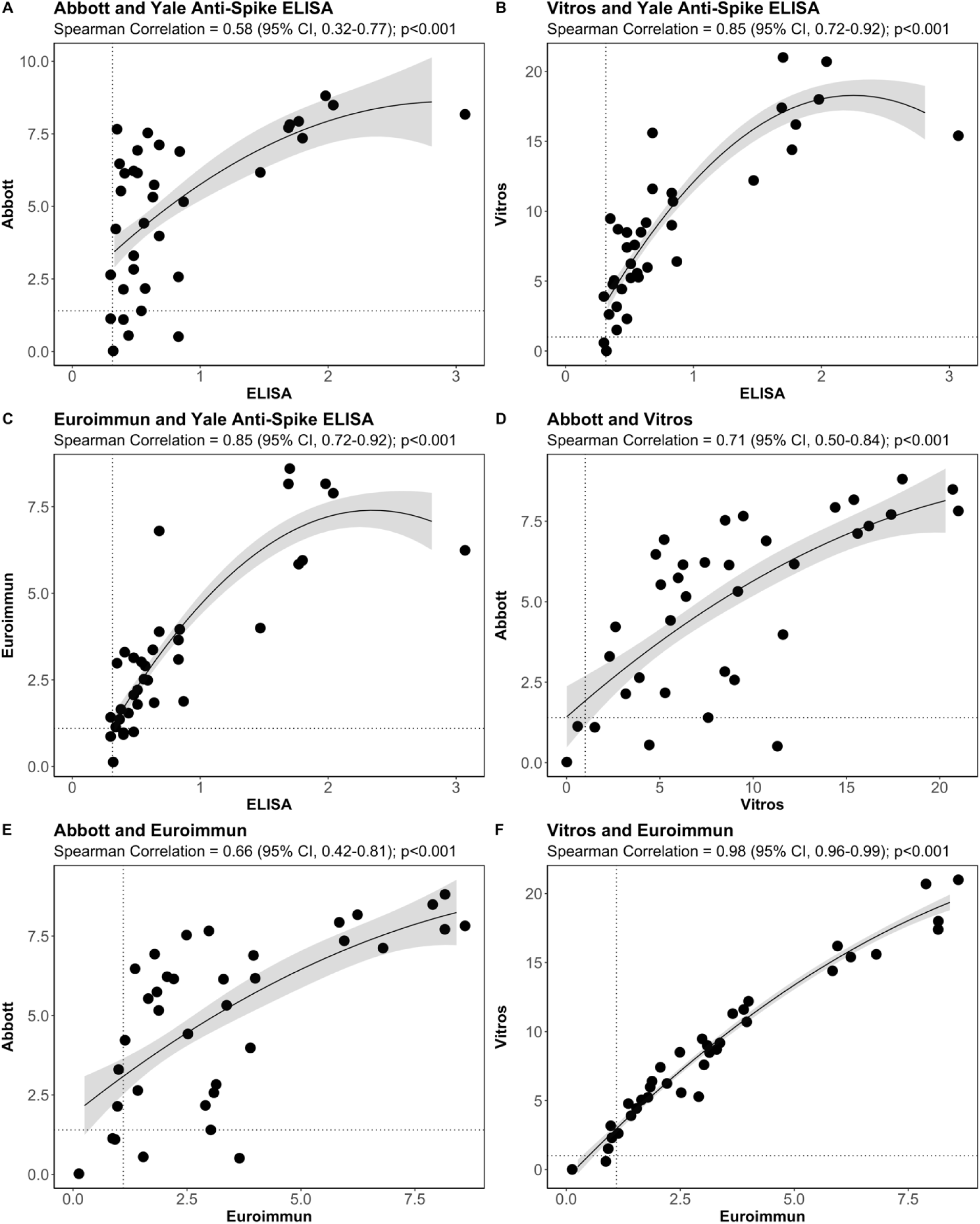
Correlations between the Yale Anti-Spike ELISA, Abbott Architect, Ortho Vitros, and Euroimmun SARS-CoV-2 IgG Assays. The dashed line represents the cutoffs used. Abbreviations: CI, confidence interval; ELISA, enzyme-linked immunosorbent assay; SARS-CoV-2, severe acute respiratory syndrome coronavirus 2.

## DISCUSSION

This study assessed the performance characteristics of four anti-SARS-CoV-2 serology tests in individuals with a prior SARS-CoV-2 infection. The sensitivities of the tests were not significantly different, and the quantitative scales of all assays were strongly correlated with Spearman coefficients greater than 0.58 and p<0.001 in all cases. While the Ortho Vitros and the Euroimmun assays had a very strong correlation with each other, and good correlation with the Yale anti-spike ELISA, the Abbott Architect assay, the only anti-nucleocapsid test, exhibited lower correlation with the three other tests in the comparison.

The 3 different FDA EUA SARS-CoV-2 IgG assays and the Yale anti-spike ELISA used in this study detect antibodies directed against different immunogenic components of SARS-CoV-2. The spike protein is the main surface glycoprotein of SARS-CoV-2 that is used to attach and enter cells,^6^ whereas the nucleocapsid is critical for viral transcription and replication. Initial studies suggested that assays using the SARS-CoV-2 nucleoprotein may have higher sensitivity.^7,8^ However, the data presented here show a qualitatively higher sensitivity with the 3 assays that use the spike glycoprotein (Ortho Vitros, Euroimmun, Yale ELISA), as compared with the Abbott Architect assay, suggesting a need for further comparative testing. Additionally, as expected, the 3 anti-spike assays had a stronger correlation among themselves as compared with the anti-nucleocapsid assay.

Given that the sensitivities of four anti-SARS-CoV-2 protein assays did not significantly differ, it may be reasonable to use any of these assays to assess the prevalence of SARS-CoV- 2 antibodies. However, recognizing differences in the correlation between these assays can help facilitate interpretation of prevalence studies using the different assays. For example, given the very strong correlation between Euroimmun and Ortho Vitros assays, it would be reasonable to aggregate seroprevalence studies that use these assays into meta-analysis of seroprevalence or a comparison over time. The weaker correlation of the Abbott Architect with the other serology assays, however, suggest caution should be used when aggregating seroprevalence rates detected by the Abbott Architect assay with those using other serologic assays. Studies focusing on assessment of seroprevalence of COVID-19 in the different populations should consider for the sensitivity of the assay used, and the likely prevalence of antibodies in the population, in order to accurately interpret the results. It is also important to know the thresholds for positivity and how tests in the indeterminate range are reported.

Our study has several limitations. Our comparative study focused on the distribution and correlation of anti-SARS-CoV-2 IgG ELISA results among individuals who had confirmed COVID-19, and we were not able to evaluate the specificities of the tests. In addition, our sample size limited our power to detect clinically meaningful differences in sensitivities. Another limitation is that we had limited information on the clinical characteristics of these individuals, which could have affected the nature of antibody response. Nevertheless, this study provides important information on the correlations between 3 commercially available serology tests, as well as the Yale anti-spike ELISA, and also information on the sensitivity of these tests, which can help guide future research on the seroprevalence of IgG antibodies to SARS-CoV-2.

In conclusion, this study shows that the 3 commercial SARS-CoV-2 IgG assays (Abbott Architect, Ortho Vitros, and Euroimmun) and the Yale anti-spike ELISA, had sensitivities that were not significantly different. Additionally, the 3 assays based on the spike protein showed the strongest correlations in detecting SARS-CoV-2 antibodies and had a weaker correlation with the serology test based on the nucleocapsid protein. As such, serologic surveillance of past SARS-CoV-2 infection should consider the individual test characteristics and the correlation between different tests when comparing or aggregating data across different populations.

## Data Availability

All data referred to in the manuscript can be made available on request.

## DISCLOSURES

Dr. Krumholz works under contract with the Centers for Medicare & Medicaid Services to support quality measurement programs; was a recipient of a research grant, through Yale, from Medtronic and the United States Food and Drug Administration to develop methods for post-market surveillance of medical devices; was a recipient of a research grant with Medtronic and is the recipient of a research grant from Johnson & Johnson, through Yale University, to support clinical trial data sharing; was a recipient of a research agreement, through Yale University, from the Shenzhen Center for Health Information for work to advance intelligent disease prevention and health promotion; collaborates with the National Center for Cardiovascular Diseases in Beijing; receives payment from the Arnold & Porter Law Firm for work related to the Sanofi clopidogrel litigation, from the Ben C. Martin Law Firm for work related to the Cook Celect IVC filter litigation, and from the Siegfried and Jensen Law Firm for work related to Vioxx litigation; chairs a Cardiac Scientific Advisory Board for UnitedHealth; was a participant/participant representative of the IBM Watson Health Life Sciences Board; is a member of the Advisory Board for Element Science, the Advisory Board for Facebook, and the Physician Advisory Board for Aetna; and is a co-founder of HugoHealth, a personal health information platform, and co-founder of Refactor Health, an enterprise healthcare artificial intelligence-augmented data management company. The other co-authors report no potential competing interests.

## FUNDING

None.

## Notes

### Author Declarations

Institutional Review Board at Yale University

